# Beyond the Apnea-Hypopnea Index: Physiological and Demographic Predictors of Excessive Daytime Sleepiness in Obstructive Sleep Apnea

**DOI:** 10.64898/2026.06.12.26355543

**Authors:** Charlize Tan, Ankit Parekh, Sajila Wickramaratne

## Abstract

Excessive daytime sleepiness (EDS) is a common but inconsistently predicted symptom of obstructive sleep apnea (OSA). OSA is typically diagnosed with polysomnography (PSG), and the current standard for severity assessment is the apnea-hypopnea index (AHI). AHI has many limitations, including its inability to explain physiological mechanisms or reflect variability in patient symptoms, such as EDS. This retrospective study aims to find physiological and demographic parameters that better predict EDS in patients with OSA and to evaluate whether these parameters outperform AHI using PSG data from the Mount Sinai Integrative Sleep Center. Clinical variables used to predict EDS included arousal index (AI), average oxygen desaturation during sleep, average heart rate during sleep, and AHI, along with demographic variables including age, sex, and BMI. Hypothesis tests, logistic regression models, and decision tree classifier models were performed on the data to discriminate sleepy from nonsleepy patients as determined by an Epworth Sleepiness Scale (ESS) score ≥ 10. AI and oxygen desaturation were found to be the most predictive physiological variables, and sex and BMI were found to be the most predictive demographic variables. The final decision tree model with these four variables outperformed the AHI in predicting EDS. These findings suggest that daytime sleepiness in OSA can be better explained by measures of apnea burden, oxygenation impairment, and patient demographics than by AHI alone, although these remain only modestly predictive. Future studies should focus on investigating more comprehensive physiological markers, multi-night sleep data, and more objective assessments of sleepiness.

## 1. Introduction

Obstructive Sleep Apnea (OSA) is a sleep disorder affecting nearly 1 billion people worldwide that is characterized by repeated airway collapse, which causes sleep fragmentation and oxygen desaturation. Risk factors include obesity, male sex, old age, and structural characteristics like large tonsils. Individuals with OSA may experience loud snoring, waking up choking, fatigue, or difficulty sleeping. It can lower patients’ quality of life and is associated with increased risks of hypertension, cardiovascular disease, diabetes, motor vehicle accidents, stroke, and neurocognitive impairment (Shahar et al., 2001; Silva et al., 2016; Tregear et al., 2009; Young et al., 1997). To diagnose OSA, patients undergo an overnight polysomnography (PSG) in a lab, which records sleep and breathing patterns through various sensors that measure respiratory effort, movement, airflow, blood oxygen, apneas/hypopneas, and more.

The standard metric for determining the severity of OSA is the apnea-hypopnea index (AHI), which counts the number of apnea and hypopnea events per hour. An AHI of 5 is the threshold for OSA, with 5-15 classified as mild OSA, 15-30 as moderate OSA, and over 30 indicating severe OSA. Studies have shown correlations between AHI and consequences of OSA, including hypertension, quality of life, and stroke (Jean-Louis et al., 2008; Malhotra et al., 2021). However, there are drawbacks to AHI as a metric. One concern is the differing definitions of apneas and hypopneas, which leads to inconsistent AHI methodology. Additionally, AHI doesn’t fully reflect the underlying physiological reasons behind cardiovascular, neurocognitive, and metabolic effects (Soori et al., 2022). It has a limited ability to explain the variability in symptoms resulting from OSA for people with the same score and to predict outcomes and response to treatment.

The most common consequence of OSA is excessive daytime sleepiness (EDS), a condition characterized by an overwhelming need to sleep during the day. It can be caused by inadequate sleep as well as sleep disorders like OSA, narcolepsy, and circadian rhythm disorders. It is also associated with psychiatric, mood, and medical disorders, as well as old age, male sex, and high BMI (Block et al., 1979). Individuals with EDS may experience sleep attacks, unintended naps, and/or difficulty waking. EDS could reduce quality of life by negatively impacting performance in professional, social, and academic settings. It also raises the risks of diabetes, cardiovascular issues, and cognitive impairment. Beyond its personal impact, EDS can also pose danger to others, as it is linked to an increased risk of motor vehicle accidents and medical errors.

In our study, EDS is defined as an Epworth Sleepiness Scale (ESS) score ≥ 10. The ESS measures general sleepiness on a scale of 0-24 by asking patients to rate their likelihood of falling asleep on a scale of 0-3 across 8 different scenarios, with a score of 10 or higher indicating sleepiness. Studies have shown that EDS defined by ESS is poorly correlated with AHI and has a weak ability to distinguish sleepy from nonsleepy patients (Kapur et al., 2005).

Despite the clinical importance of EDS, existing metrics do not adequately predict sleepiness. This study aims to evaluate whether other physiological and demographic variables (AHI, arousal index, average oxygen desaturation, average heart rate, BMI, sex, and age) derived from PSG, or a combination of parameters, better predict EDS than AHI alone.

## 2. Methods

### 2.1. Population

This study included 8518 patients from the Nexus360 database who underwent on-site polysomnography (PSG) evaluation at the Mount Sinai Integrative Sleep Center. An IRB was obtained, and all patients provided informed consent for the anonymous use of their data under approved protocol.

Collected data included AHI, arousal index (defined as the number of electroencephalogram (EEG) arousals per hour of sleep), average oxygen desaturation (defined as the average drop in blood oxygen), average heart rate, BMI (calculated from height and weight), sex, and age. The study excluded ESS = 0 or NA (n = 2869) and any incomplete cases for the eight variables measured (n = 2969). Among participants in the study (n = 2680), 1153 (43%) were classified as sleepy (ESS ≥ 10), and 1527 (57%) were classified as nonsleepy (ESS<10). Patients were 18-90 years old. 1326 (49.5%) were female, and 1354 (50.5%) were male. BMI ranged from 15.21 to 49.95.

### 2.2. Statistical Analysis

Statistics were performed in R (version 4.5.1), using packages ggplot2 (plots), gtsummary (tables), modelsummary (tables), stargazer (logistic regression tables), pROC (AUROC metric), and performance (check assumptions). Normally distributed variables were reported as mean ± standard deviation and analyzed using independent sample t-tests, and non-normally distributed variables were reported as median (min, max) and analyzed using the Wilcoxon rank-sum test. A p-value < 0.05 was considered statistically significant. Univariate linear regression was used to identify correlates of daytime sleepiness, with the ESS score as the dependent variable. Univariate logistic regression analyses were then performed separately for physiological variables, with EDS defined as ESS ≥ 10. Demographic variables were included in a second round of analysis, and the final logistic regression model incorporated only statistically significant parameters. Decision tree classifiers were also used to analyze predictors of EDS, in which the algorithm used physiological and demographic parameters to split the data and subsequent subsets of data into terminal nodes representing a categorization of an observation as either sleepy or nonsleepy group. Independent variables in the decision tree were chosen based on performance in linear and logistic regression models, and the final model included only statistically significant variables. Data had a 70-30 split for training and testing. Parameters for the final decision tree model were chosen to minimize overfitting and underfitting. They were set as maxdepth = 7, cp = 0.008, and minsplit = 75.

Linear regressions were analyzed using their R^2^ value. Logistic regression and decision tree classifier models were evaluated using the Area Under the Curve (AUC), which combines the true positive and false positive rates. In this study, AUC was used to assess the model’s ability to predict EDS based on clinical variables. Trees were grown in packages caret, rpart, and randomforest. The confusion matrix was also used to assess models by looking at the distribution of classifications across the test set.

## 3. Results

To evaluate the characteristics most predictive of daytime sleepiness, we analyzed AHI, AI, desaturation, heart rate, and demographics (age, sex, BMI) using both linear and logistic regression. Additionally, we separated the analysis into key variables, including and excluding demographics. Table I shows the center and spread of each variable separated into sleepy (1) and nonsleepy (0) groups. These variables demonstrate a wide range of variability amongst patients, consistent with the heterogeneous presentation of OSA. There were more nonsleepy people included in the study than sleepy, and the sample is balanced across sex. Hypothesis tests were run on each variable, and p-values are included. There were significant differences between the sleepy and nonsleepy groups only for AHI, AI, and age. Of note, the median AHI, AI, and age are higher for the nonsleepy group, which contrasts with the expectation of higher values for the sleepy group.

**Table I:**
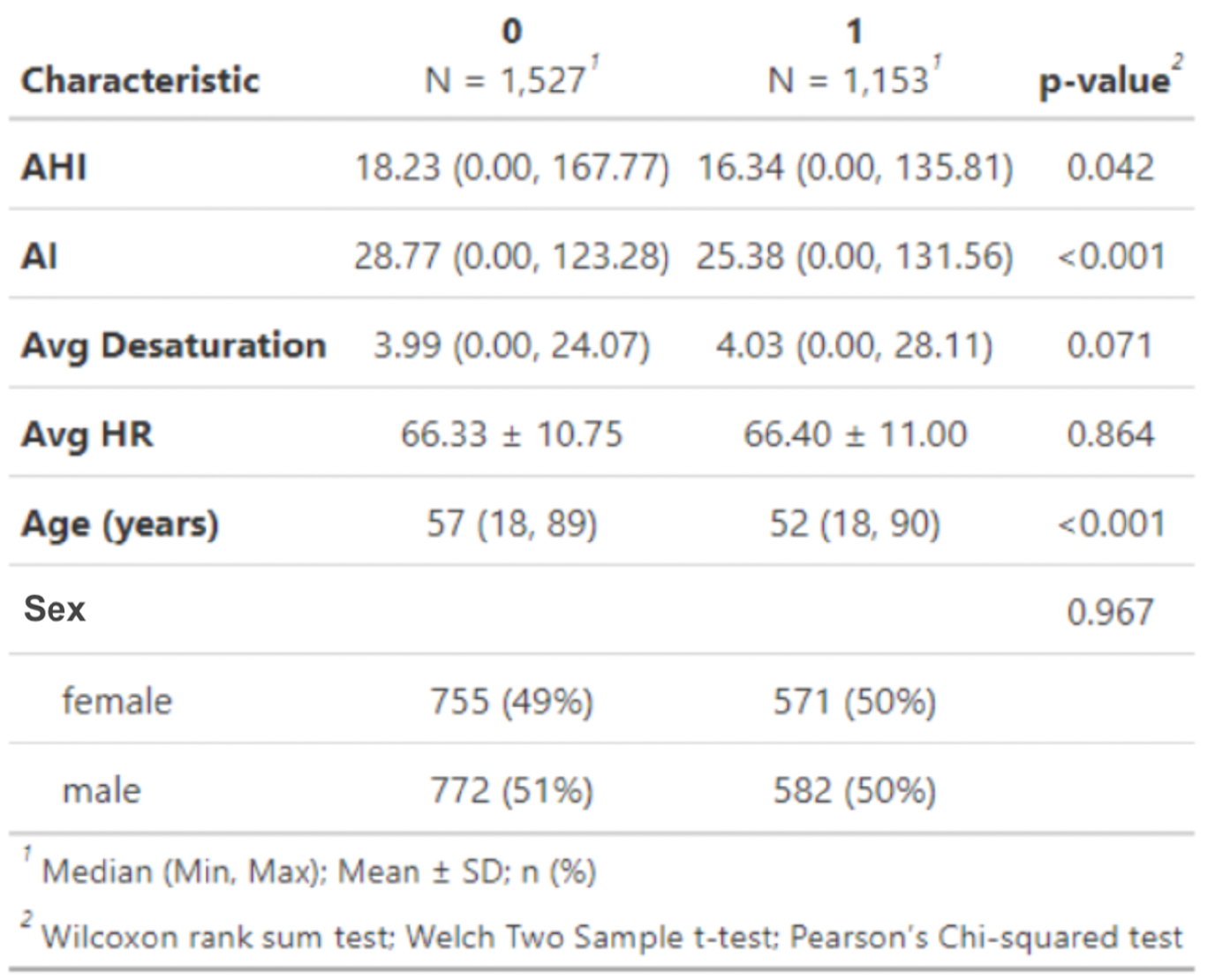
Center and Spread of Patient Variables across Sleepy (1) and Nonsleepy (0) Groups Caption: Center and spread of patient variables included in the study compared across sleepy and nonsleepy groups using the Wilcoxon rank sum test, Welch two-sample t-test, and Pearson’s Chi-squared test. AHI = Apnea-Hypopnea Index AI = Arousal Index HR = Heart Rate

### 3.1. Logistic Regression Analysis

To determine which physiological metrics were most strongly associated with daytime sleepiness, a series of logistic regression models was first performed without demographic variables. First, models examined AHI, AI, desaturation, and HR as isolated predictors of sleepiness, and an additional model evaluated a combined predictor using AI and desaturation. The analyses were performed in R, and the AUC and p-values were used to evaluate the models, which are common metrics for binary classification. Significant variables, AI and desaturation, were then combined into one model (Model 5) and analyzed with logistic regression to test for a higher AUC. Table II shows that, among the individual variables, AI had the highest AUC value of 0.544. Model 5, which included AI and desaturation, had an even higher AUC of 0.551, indicating that it was the most predictive model. Despite the increase in AUC, it is important to note that the value for each variable ranges from 0.5 to 0.551, indicating little to no discriminatory power, thus providing low predictiveness of daytime sleepiness.

**Table II:**
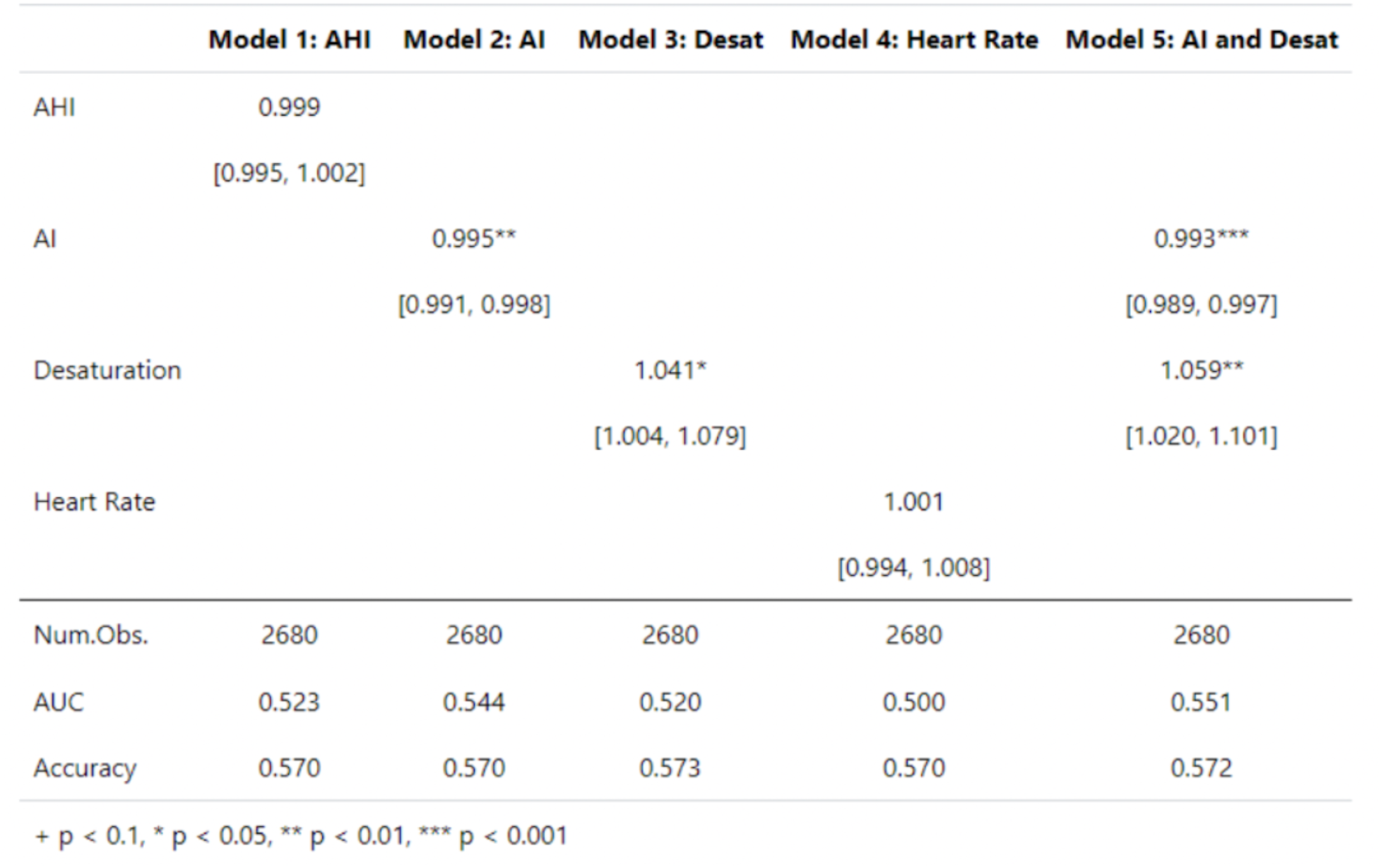
Logistic Regression Analysis showing the Combined Model of AI and Desaturation as the Most Predictive Indicator of Daytime Sleepiness Caption: Logistic regression models of AHI, AI, desaturation, heart rate, and AI + desaturation. In the top half of the figure, the top number shown is the odds ratio, and the number in brackets is the confidence interval.

A second set of logistic regression models was fitted, utilizing both demographic variables and physiological predictors. These models incorporated age and sex, along with AHI, AI, desaturation, and heart rate. After adjusting for demographics, each model’s performance improved, and the combined AI and desaturation model remained the most predictive of daytime sleepiness. This pattern supports the expectation that deeper desaturation episodes and more frequent arousals, both of which cause sleep fragmentation, lead to more daytime sleepiness. As shown in Table III, AUC values ranged from 0.576 to 0.583, indicating modest predictive power but an improvement over previous models. These findings reinforce that sleepiness in OSA is multifactorial and dependent on both physiological and demographic variables.

**Table III:** Logistic Regression Analysis showing the Combined Model of AI and Desaturation with Demographics as the Most Predictive Indicator of Daytime Sleepiness Caption: Logistic regression models of AHI, AI, desaturation, heart rate, and AI + desaturation with demographic variables added. In the top half of the figure, the top number shown is the odds ratio, and the number in brackets is the confidence interval.

|  | Model 1: AHI with demographics | Model 2: AI with demographics | Model 3: Desat with demographics | Model 4: Heart Rate with demographics | Model 5: AI and Desat with demographics |
| --- | --- | --- | --- | --- | --- |
| AHI | 1.001<br>[0.998, 1.005] |  |  |  |  |
| AI |  | 0.998<br>[0.994, 1.001] |  |  | 0.996*<br>[0.992, 1.000] |
| Desaturation |  |  | 1.059**<br>[1.020, 1.099] |  | 1.068***<br>[1.028, 1.111] |
| Heart Rate |  |  |  | 0.997<br>[0.990, 1.004] |  |
| Age | 0.983***<br>[0.978, 0.988] | 0.985***<br>[0.980, 0.990] | 0.983***<br>[0.978, 0.988] | 0.984***<br>[0.979, 0.988] | 0.984***<br>[0.979, 0.989] |
| Sex | 1.010<br>[0.864, 1.180] | 1.027<br>[0.880, 1.200] | 0.994<br>[0.851, 1.161] | 1.002<br>[0.856, 1.175] | 1.008<br>[0.863, 1.179] |
| Num.Obs. | 2680 | 2680 | 2680 | 2680 | 2680 |
| AUC | 0.576 | 0.577 | 0.580 | 0.576 | 0.583 |
| Accuracy | 0.574 | 0.573 | 0.578 | 0.574 | 0.578 |

### 3.2. Decision Tree

To test for the possibility of a higher AUC, a decision tree classifier was constructed using all physiological and demographic predictors, including AHI, AI, desaturation, heart rate, BMI, age, and sex. The visualization of the final model shows that its algorithm identified desaturation and AI as the two most predictive physiological variables, and age and BMI as the two most important demographic predictors of daytime sleepiness (Figure 1A). After applying the decision tree classifier to the training and testing data, the AUC increased from the logistic regression models to 0.598, indicating that daytime sleepiness is best predicted by a combination of AI, desaturation, age, and BMI rather than AHI alone (Figure 1B).

**Figure 1:**
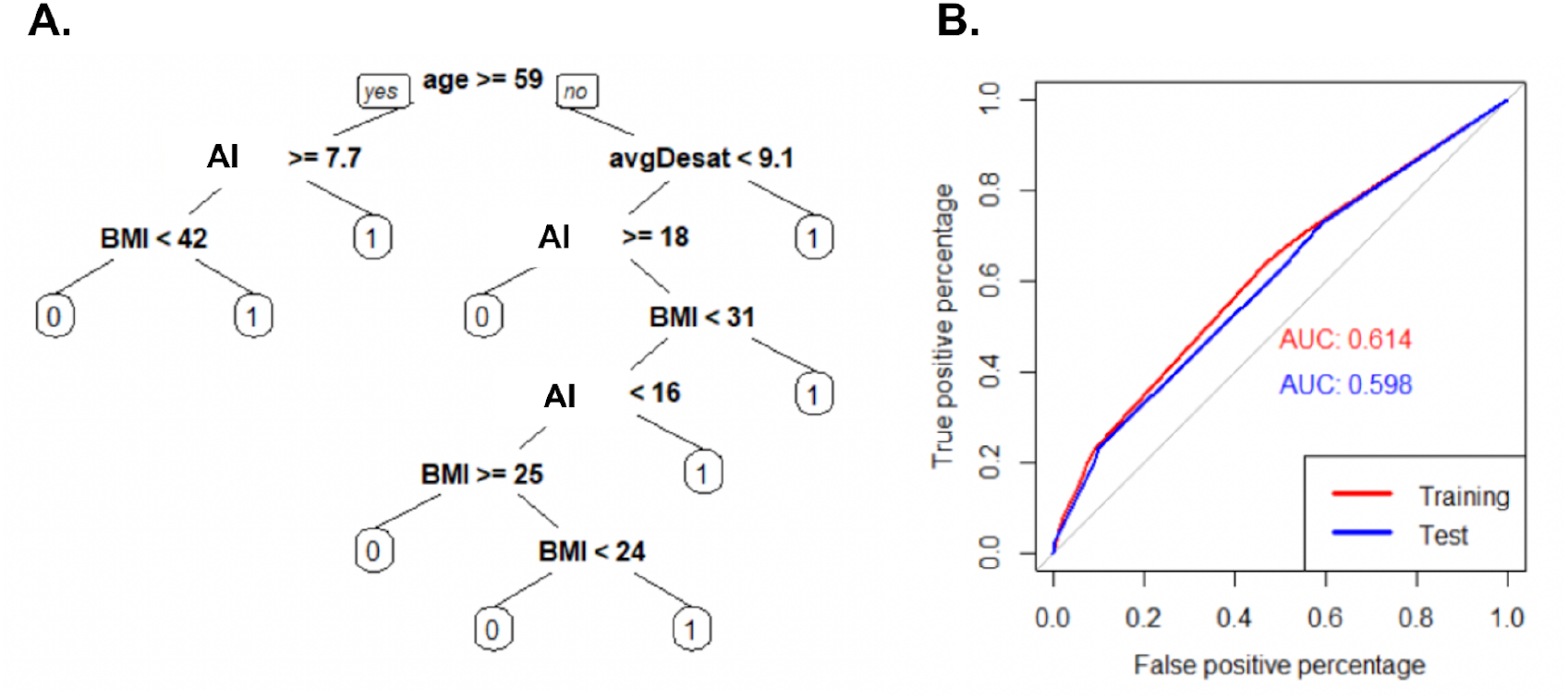
Decision Tree Model and AUC Graph Indicating AI, Desaturation, Age, and BMI as the Most Predictive Variables of Daytime Sleepines. Caption: A. Decision tree classifier splitting patients into nonsleepy (0) and sleepy (1) groups based on physiological and demographic variables. B. AUC graph showing the true and false positive percentages of training and testing data for the decision tree classifier.

## 4. Discussion

This study sought to identify physiological and demographic determinants that influence daytime sleepiness in patients with OSA. Findings from logistic regression and decision tree analyses indicated that four variables, AI, oxygen desaturation, age, and BMI were most consistently predictive of EDS. In contrast, AHI, the most commonly used clinical parameter, demonstrated weaker and less predictive values. These findings highlight the multifactorial nature of daytime sleepiness in OSA and suggest that AHI alone is not indicative of an accurate level of daytime sleepiness.

Logistic regression models found AI and oxygen desaturation to be the two strongest physiological predictors of sleepiness, outperforming AHI and heart rate, both individually and in combination. This finding supports the notion that oxygen impairment and the frequency of complete obstructive events, rather than the combined burden of apneas and hypopneas, may better capture the sleep disturbances and hypoxic stressors that link to neurocognitive outcomes like EDS. This pattern suggests a physiological relationship that deeper desaturation episodes can exacerbate sympathetic activation, oxidative stress, and sleep fragmentation, all of which contribute to daytime sleepiness. It is important to note that all models without demographic variables demonstrated minimal discriminatory power (AUC between 0.50 and 0.55). This finding underscores that physiological metrics alone may not fully capture the complex and chronic processes underlying daytime sleepiness.

After including age and sex into the regression models, predictive performance improved (AUC between 0.576 and 0.583). Among these, age was a significant demographic contributor, which was consistent with known age-related decline in arousal threshold, respiratory stability, and the structural integrity of the upper airway (Eikermann et al., 2007). Although not part of the logistic regression models, BMI was identified by the decision tree as another key factor influencing sleepiness. The decision tree identified AI, desaturation, BMI, and age as the most predictive of EDS and had the highest AUC of 0.598 on testing data. BMI is strongly linked to OSA pathophysiology through increased upper airway collapsibility and greater average neck circumference, which may exacerbate the frequency of upper airway events. These results support a growing body of evidence indicating that demographic variables like age-related neurophysiological changes and obesity-related mechanical factors influence daytime sleepiness in OSA (Panossian et al., 2012).

### 4.1 Limitations

The study has a few limitations that may have influenced the accuracy and quality of the results. The primary limitation was the use of ESS to measure the level of sleepiness in the participants. The ESS is a self-administered test comprising eight questions that require respondents to evaluate their sleepiness on a four-point scale. Participants are asked to quantitatively evaluate their subjective sleepiness in a categorical manner; however, a continuous or multidimensional scale would probably be more representative. More objective methods such as the Multiple Sleep Latency Test would yield more results, but since it is more complicated and time-consuming, it would be difficult to include a large cohort. Second, the study used only a subset of standard PSG variables, which may not be able to fully reflect the nuanced and possibly more accurate measurements of EDS. The inclusion of variables such as the duration of respiratory events, oxygen desaturation, ventilatory burden, and heart rate variability would improve this (Kainulainen, 2019; Ma et al., 2021; Parekh, 2023; Ucak, 2024; Varghese, 2022). Thirdly, this study did not consider confounding factors like medication use, insomnia, psychiatric disorders, and circadian rhythm disorders.

Besides better variables, multi-night sleep data would provide a better account of any night-to-night sleep variation that a single-night PSG would not be able to capture accurately. Other machine learning methods such as XGBoost and RandomForest might also be employed to explore the multifactorial aspects of sleep apnea and daytime sleepiness and take into account more comprehensive demographics and clinical variables. Finally, a larger and more balanced dataset would enhance model performance and generalizability.

## 5. Conclusion

This study illustrates that AHI alone is less predictive of EDS in patients with OSA than the combined use of physiological and demographic variables, in particular oxygen desaturation, arousal index, age, and BMI. These results support that the severity of OSA cannot be fully accounted for by the frequency of apneas and hypopneas alone and that the causes of daytime sleepiness are, to a greater extent, related to sleep fragmentation, hypoxic burden, and patient-specific factors.

Although the overall predictive performance of all models was limited, the consistently superior performance of parameters other than AHI suggests that using AHI as a universal marker of clinical severity is likely to hide important physiology. The integration of arousal burden, oxygenation impairment, and demographic context measures may pave the way for a more thorough characterization of OSA and its consequences linked to daytime sleepiness.

Future research should focus on identifying more comprehensive physiological markers, incorporating multi-night sleep data, and employing objective measures of sleepiness. Going beyond the AHI-based classification may not only benefit risk stratification but also clinical decision-making for OSA patients, particularly those whose symptoms are not consistent with traditional severity metrics.

## Data Availability

The data analyzed in this study were obtained from the Icahn School of Medicine at Mount Sinai under approved research protocols and are not publicly available due to institutional and participant privacy restrictions.

## Bibliography

Block, A. J., Boysen, P. G., Wynne, J. W., & Hunt, L. A. (1979). Sleep apnea, hypopnea and oxygen desaturation in normal subjects. A strong male predominance. The New England journal of medicine, 300(10), 513–517. 10.1056/NEJM197903083001001

Eikermann, M., Jordan, A. S., Chamberlin, N. L., Gautam, S., Wellman, A., Lo, Y. L., White, D. P., & Malhotra, A. (2007). The influence of aging on pharyngeal collapsibility during sleep. Chest, 131(6), 1702–1709. 10.1378/chest.06-2653

Jean-Louis, G., Zizi, F., Clark, L. T., Brown, C. D., & McFarlane, S. I. (2008). Obstructive sleep apnea and cardiovascular disease: role of the metabolic syndrome and its components. Journal of clinical sleep medicine : JCSM : official publication of the American Academy of Sleep Medicine, 4(3), 261–272.

Kainulainen, S., Töyräs, J., Oksenberg, A., Korkalainen, H., Sefa, S., Kulkas, A., & Leppänen, T. (2019). Severity of Desaturations Reflects OSA-Related Daytime Sleepiness Better Than AHI. Journal of clinical sleep medicine : JCSM : official publication of the American Academy of Sleep Medicine, 15(8), 1135–1142. 10.5664/jcsm.7806

Kapur, V. K., Baldwin, C. M., Resnick, H. E., Gottlieb, D. J., & Nieto, F. J. (2005). Sleepiness in patients with moderate to severe sleep-disordered breathing. Sleep, 28(4), 472–477. 10.1093/sleep/28.4.472

Koutsourelakis, I., Perraki, E., Bonakis, A., Vagiakis, E., Roussos, C. & Zakynthinos, S. (2008) Determinants of subjective sleepiness in suspected obstructive sleep apnoea. Journal of Sleep Research, 17: 437–443. 10.1111/j.1365-2869.2008.00663.x

Leng, P. H., Low, S. Y., Hsu, A., & Chong, S. F. (2003). The clinical predictors of sleepiness correlated with the multiple sleep latency test in an Asian Singapore population. Sleep, 26(7), 878–881. 10.1093/sleep/26.7.878

Ma, C., Zhang, Y., Liu, J. et al. A novel parameter is better than the AHI to assess nocturnal hypoxaemia and excessive daytime sleepiness in obstructive sleep apnoea. Sci Rep 11, 4702 (2021). 10.1038/s41598-021-84239-0

Malhotra, A., Ayappa, I., Ayas, N., Collop, N., Kirsch, D., Mcardle, N., Mehra, R., Pack, A. I., Punjabi, N., White, D. P., & Gottlieb, D. J. (2021). Metrics of sleep apnea severity: beyond the apnea-hypopnea index. Sleep, 44(7), zsab030. 10.1093/sleep/zsab030

Panossian, L. A., & Veasey, S. C. (2012). Daytime sleepiness in obesity: mechanisms beyond obstructive sleep apnea--a review. Sleep, 35(5), 605–615. 10.5665/sleep.1812

Parekh, A., Kam, K., Wickramaratne, S., Tolbert, T. M., Varga, A., Osorio, R., … & Rapoport, D. M. (2023). Ventilatory burden as a measure of obstructive sleep apnea severity is predictive of cardiovascular and all-cause mortality. American Journal of Respiratory and Critical Care Medicine, 208(11), 1216–1226.

Shahar, E., Whitney, C. W., Redline, S., Lee, E. T., Newman, A. B., Nieto, F. J., O’Connor, G. T., Boland, L. L., Schwartz, J. E., & Samet, J. M. (2001). Sleep-disordered breathing and cardiovascular disease: cross-sectional results of the Sleep Heart Health Study. American journal of respiratory and critical care medicine, 163(1), 19–25. 10.1164/ajrccm.163.1.2001008

Silva, G. E., Goodwin, J. L., Vana, K. D., & Quan, S. F. (2016). Obstructive Sleep Apnea and Quality of Life: Comparison of the SAQLI, FOSQ, and SF-36 Questionnaires. Southwest journal of pulmonary & critical care, 13(3), 137–149. 10.13175/swjpcc082-16

Soori, R., Baikunje, N., D’sa, I., Bhushan, N., Nagabhushana, B., & Hosmane, G. B. (2022). Pitfalls of AHI system of severity grading in obstructive sleep apnoea. Sleep science (Sao Paulo, Brazil), 15(Spec 1), 285–288. 10.5935/1984-0063.20220001

Tregear, S., Reston, J., Schoelles, K., & Phillips, B. (2009). Obstructive sleep apnea and risk of motor vehicle crash: systematic review and meta-analysis. Journal of clinical sleep medicine : JCSM : official publication of the American Academy of Sleep Medicine, 5(6), 573–581

Ucak, S., Dissanayake, H. U., de Chazal, P., Bin, Y. S., Sutherland, K., Setionago, B., … & Cistulli, P. A. (2024). Heart rate variability analysis in obstructive sleep apnea patients with daytime sleepiness. Sleep, 47(6), zsae075

Varghese, L., Rebekah, G., Oliver, A., & Kurien, R. (2022). Oxygen desaturation index as alternative parameter in screening patients with severe obstructive sleep apnea. Sleep science, 15(S 01), 224–228.

Young, T., Peppard, P., Palta, M., Hla, K. M., Finn, L., Morgan, B., & Skatrud, J. (1997). Population-based study of sleep-disordered breathing as a risk factor for hypertension. Archives of internal medicine, 157(15), 1746–1752

